# The effect of early-stage public health policies on the transmission of COVID-19 in South American countries

**DOI:** 10.1101/2020.08.09.20149286

**Authors:** Bryan Valcarcel, Jose L. Avilez, J. Smith Torres-Roman, Julio A. Poterico, Janina Bazalar-Palacios, Carlo La Vecchia

## Abstract

**Objectives:** The analysis of transmission dynamics is crucial to determine whether mitigation or suppression measures reduce the spread of Coronavirus disease 2019 (COVID-19). This study sought to estimate the basic (*R_0_*) and time-varying (*R_t_*) reproduction number of COVID-19 and contrast the public health measures for ten South American countries.

**Methods:** Data was obtained from the European Centre for Disease Prevention and Control. Country-specific *R_0_* estimates during the first two weeks of the outbreak and *R_t_* estimates after 90 days were estimated.

**Results:** Countries used a combination of isolation, physical distancing, quarantine, and community-wide containment measures to staunch the spread of COVID-19 at different points in time. *R_0_* ranged from 1.52 (95% confidence interval: 1.13-1.99) in Venezuela to 3.83 (3.04-4.75) in Chile, whereas *R_t_* after 90 days ranged from 0.71 (95% credible interval: 0.39-1.05) in Uruguay to 1.20 (1.19-1.20) in Brazil. Different *R_0_* and *R_t_* values may be related to the testing capacity of each country.

**Conclusion:** *R_0_* in the early phase of the outbreak varied across the South American countries. The public health measures adopted in the initial period of the pandemic appear to have reduced *R_t_* over time in each country, albeit to different levels.

## INTRODUCTION

Coronavirus disease 2019 (COVID-19) is an emerging respiratory infectious disease caused by severe acute respiratory syndrome coronavirus 2 (SARS-CoV-2). COVID-19 was first detected in December 2019 in Wuhan, China, and has caused serious public health concerns worldwide (1). Several public health measures—physical isolation; temporary closure of territory borders, academic institutions, and public places; and quarantine—have been used in an effort to reduce the impact of the COVID-19 outbreak. The United Nations highlights that the COVID-19 pandemic, in combination with the profound socioeconomic inequalities in SA, will push over 45 million people into poor conditions and 28 million in extremely poor conditions (2). Compounded with the fragmented and ill-prepared public health systems of South American (SA), COVID-19 could have a comparatively harsher effect in the region, relative to countries with robust health systems (3).

Although the first case of COVID-19 in SA was detected on March 26 in Brazil (4), few SA modeling studies have emerged since (5). Given the rapid spread of SARS-CoV-2 in this region, understanding the dynamics of disease transmission is key to guiding the implementation of necessary prevention and control measures. One parameter, the basic reproduction number (*R*_0_), aids to fulfill this purpose by estimating the number of secondary cases arising from exposure to an infected person in the absence of epidemic or pandemic containment measures (6). Another metric, the time-varying reproduction number (R_t_), is useful in monitoring the transmissibility of SARS-CoV-2 over time and to assess the adequacy of current control efforts. *R_t_* estimates the expected number of secondary infections from an infected individual at time *t* (7, 8). The serial interval is another key variable to estimate *R_0_* and *R_t_*; it measures the time elapsed between symptomatic cases in a chain of transmission. Determining the probability distribution of the serial interval of SARS-CoV-2 and using that distribution to estimate *R_0_* and *R_t_* is crucial in assessing the rate at which the COVID-19 pandemic expands. Knowing the person-to-person transmission rate helps policymakers understand whether mitigation and suppression measures are effective and when to adopt more or less stringent measures (9, 10).

Given the importance of assessing the public health interventions to monitor their effectiveness, we estimated both reproduction numbers *(R_0_* and *R_t_*) to identify the impact of the early-stage public health interventions in South America.

## MATERIAL AND METHODS

### Data sources

We used the COVID-19 database from the European Centre for Disease Prevention and Control (ECDC) (11). The database is publicly available and contains the worldwide geographic distribution of COVID-19 cases. The database is updated daily and provides counts for new cases and deaths by date and country. We extracted the data from the ECDC database for the following ten SA countries: Argentina, Bolivia, Brazil, Chile, Colombia, Ecuador, Paraguay, Peru, Uruguay, and Venezuela. Early-stage containment and mitigation decrees issued by each country against COVID-19 were found in the corresponding official government webpages.

### Variables

The following binary variables were recorded for each country for qualitative analysis: *isolation*, separation of confirmed cases with COVID-19 in a healthcare facility or their home; *quarantine*, social restriction and home containment of persons with suspected or known contact with a patient with COVID-19, or individuals with a travel history to Europe or Asia; *physical distancing*, group of measures related to the prevention of mass gatherings, closure of academic institutions, and cancellation of social and public events; *community-wide containment*, mandatory isolation of every citizen of the country in their home, with permission only to acquire life supplies (i.e. food or water) in restricted hours of the day (12).

### Basic reproductive number (R_0_) and time-varying reproductive number (R_t_)

We estimated *R_0_* using data for the first two weeks after the first laboratory-confirmed case of SARS-CoV-2 in each country. Since *R_0_* is the reproductive rate *sans* intervention measures, it should be estimated during the exponential phase of the pandemic; otherwise, *R_0_* would be underestimated. Thus, the two-week estimation time frame was selected for two reasons: (i) 14 days is regarded as the maximum time after exposure for symptoms to develop, and (ii) it is sensible to assume that the pandemic will be in its exponential phase for the first two weeks after the first case is detected. We specified the SARS-CoV-2 serial interval as a gamma-distributed random variable with a mean serial time of 3.96 days and a standard deviation of 4.75 days, based on a contact tracing study conducted by the Center for Disease Control and Prevention (CDC) (13). *R_0_* was estimated with the maximum likelihood method described by White and Pagano (14).

We estimated *R_t_* from the time series of daily incidence of SARS-CoV-2 in each country, using the novel methods described by Thompson et al. and setting the same gamma distribution specification for the serial interval as we did for *R_0_* (15). We chose a five-day moving window to elucidate the time trend exhibited by *R_t_*. A previous study in China used a ten-day moving window to report *R_t_*(16); however, they specified a mean serial time of 7.5 days, a mean higher by a factor of two with respect to the CDC contact tracing study. We selected a narrower window to account for the faster spreading dynamics we specify in our models and estimate *R_t_* after 90 days for each country. The mean difference between *R_0_* and *R_t_* were estimated to describe the change of both numbers. Sensitivity analysis included the estimation of *R_0_* up to the first week of the first reported case in each country, based on a follow-up study of 1 000 COVID-19 cases (17).

### Data analysis

The statistical analysis was conducted using the R version 3.6.2 software. First, we describe the public health measures taken by SA countries and calculated the cumulative number of cases according to the two periods. Thereafter, the “R0” package was used to compute the basic reproductive number. To estimate both *R_0_* and *R_t_*, we employed a gamma-distributed serial interval with a mean serial interval of 3.96 ± (standard deviation) 4.75 days. Estimation of *R_0_* was carried out via the maximum likelihood method, and 95% confidence intervals (95% CI) were computed for the *R_0_* value of each country. We used the “EpiEstim” package to compute the time-varying reproductive number. Country-wise time series for *R_t_*, with 95% credible intervals (95% CrI), were plotted. The code is freely available at https://github.com/jlavileze/covidsa.

### Ethics

The project was revised by the Investigation Review Board of the Scientific University of the South. Given that the investigation is a secondary analysis of a database, it received an exempt category and was approved for its development.

## RESULTS

Table 1 shows the public health measures adopted by SA governments against COVID-19. All SA countries adopted isolation, quarantine, and physical distancing measures. However, the period between the first laboratory-confirmed case and the implementation of public health measures differed across SA nations. Seven countries, Argentina, Bolivia, Colombia, Paraguay, Peru, Uruguay, and Venezuela enacted their control policies in ≤7 days after their first case detection. Uruguay enacted the public health measures on the day they detected their first case. In Brazil, 19 days elapsed between detection and implementation of control measures.

**Table 1.** Status of the mitigation and suppression measures of South American countries.

| Countries | Isolation, quarantine, and social distancing |  | Community-wide containment |  |
| --- | --- | --- | --- | --- |
|  | Status | Days after 1<sup>st</sup> case | Status | Days after 1<sup>st</sup> case |
| Brazil | Yes | 19 | Yes<sup>a</sup> | 25 |
| Chile | Yes | 14 | Yes<sup>a</sup> | 17 |
| Ecuador | Yes | 14 | Yes | 17 |
| Argentina | Yes | 7 | Yes | 17 |
| Peru | Yes | 5 | Yes | 9 |
| Colombia | Yes | 6 | Yes | 18 |
| Paraguay | Yes | 3 | Yes | 9 |
| Bolivia | Yes | 6 | Yes | 7 |
| Uruguay | Yes | 0 | No | - |
| Venezuela<sup>b</sup> | Yes | 3 | Yes | 3 |

a Chile and Brazil developed a selective community-wide containment with different dates for specific provinces.
b Venezuela started the mitigation measures (isolation, quarantine, social distancing, and community-wide containment) on the same date.

The time to implement a community-wide containment strategy also varied between SA countries. Two countries, Bolivia and Venezuela, issued their mitigation policies in ≤7 days after detecting their first COVID-19 cases; Paraguay and Peru did so between 7-14 days after detection; and Argentina, Chile, Colombia, and Ecuador all took over 14 days (Table 1).

Moreover, Colombia (12 days) and Argentina (10 days) had the longest period between their first measures (isolation, physical distancing, and quarantine) and community-wide containment efforts. Chile implemented a stepwise selective community containment, moving one province at a time.

Table 2 shows *R_0_* and *R_t_* estimates for each country and the corresponding cumulative number of cases. During the early phase of the pandemic, *R_0_* ranged from 1.52 to 3.83. An *R_0_* of 3.83 means that, on average, each infected person will transmit the virus to roughly 4 different individuals. Brazil, Chile, and Ecuador had the highest *R_0_*; these three countries enacted their first mitigation measures ≥14 days from the initial detection of COVID-19 patients in their territory. For the countries that implemented mitigation measures 5-7 days after detection, the *R_0_* ranged from 1.60 to 2.95, while for the countries that acted within 3 days, it ranged from 1.52 to 1.74. The sensitivity analysis showed similar results; Brazil was the country with the highest *R_0_* after seven days of the first case detection. Moreover, the seven-day *R_0_* estimation ranged from 1.39 to 3.48 for countries with mitigation measures between 5-7 days and from 1.17 to 2.29 in nations that enacted measures in <3 days after their first COVID-19 case. Furthermore, all SA countries decreased their *R_t_* over time, with a mean difference ranging from 0.61 in Bolivia to 2.80 in Ecuador (Table 2 and Figure 1). At the latest assessment, *R_t_* ranged from 0.99 to 1.13 in all the countries studied. Although the point estimates for *R_t_* were below one in Bolivia and Uruguay, their 95% credible intervals are compatible with a still-growing pandemic (Figure 1 and Table 2).

**Figure 1.**
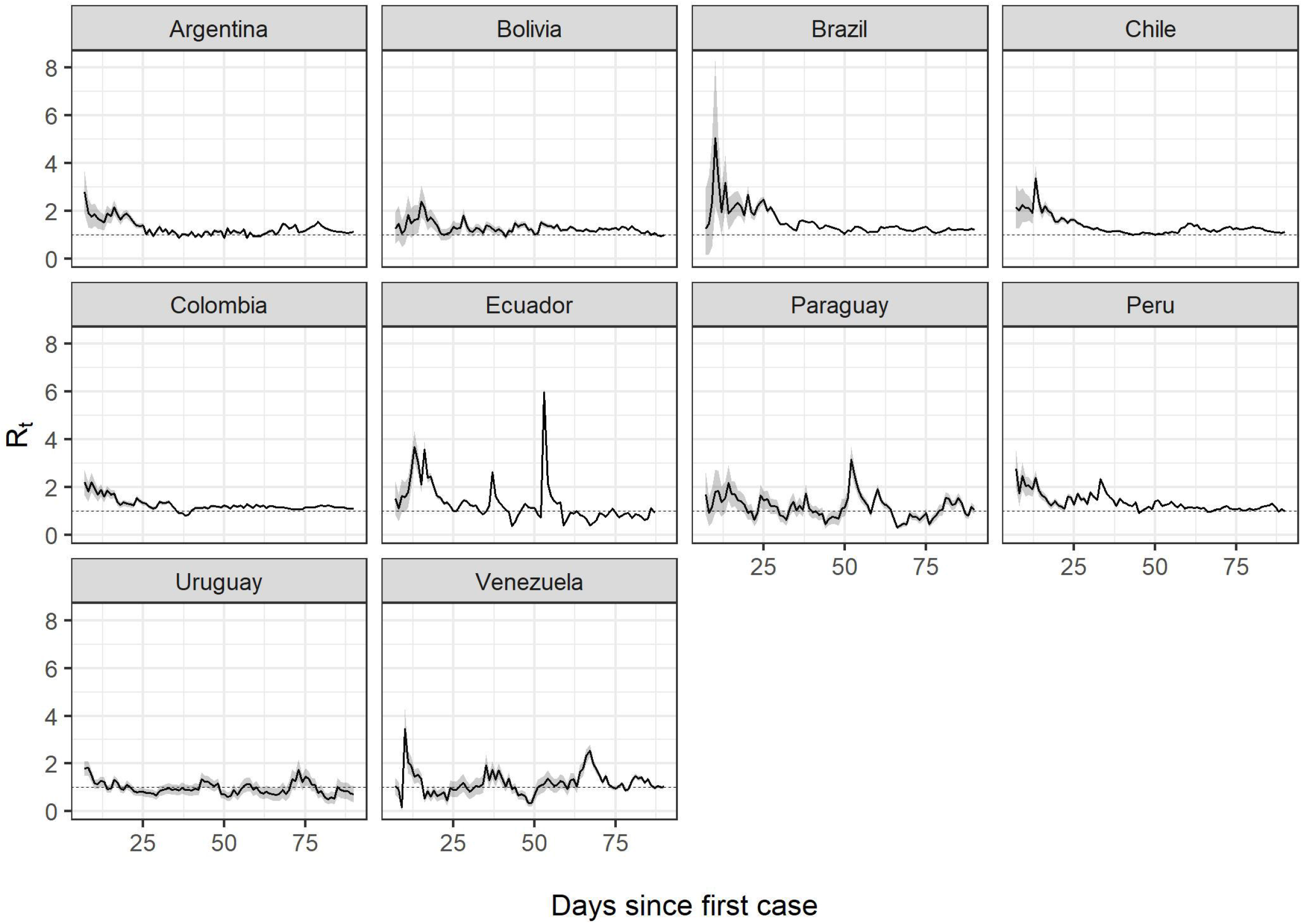
Progression of the time-varying reproduction number (*R_t_*) since the first case report in each country

**Table 2.** The basic and time-varying reproductive numbers in South American countries.

| Countries | $R_0$ after 14 days | | | $R_t$ after 90 days | | | Mean difference |
| --- | --- | --- | --- | --- | --- | --- | --- |
| | Cumulative cases | $R_0$ | 95% CI | Cumulative cases | $R_t$ | 95% CrI | |
| Argentina | 128 | 2.05 | 1.52-2.68 | 19 255 | 1.13 | 1.09-1.16 | 0.92 |
| Bolivia | 39 | 1.60 | 0.92-2.55 | 14 644 | 0.99 | 0.95-1.02 | 0.61 |
| Brazil | 25 | 3.40 | 1.82-5.70 | 363 211 | 1.2 | 1.19-1.21 | 2.20 |
| Chile | 201 | 3.83 | 3.04-4.75 | 105 159 | 1.12 | 1.11-1.14 | 2.71 |
| Colombia | 306 | 2.05 | 1.70-2.45 | 39 236 | 1.10 | 1.08-1.12 | 0.95 |
| Ecuador | 168 | 3.72 | 2.82-4.80 | 40 414 | 0.92 | 0.88-0.96 | 2.80 |
| Paraguay | 37 | 1.74 | 0.96-2.86 | 1 145 | 1.06 | 0.89-1.24 | 0.68 |
| Peru | 263 | 2.94 | 2.44-3.51 | 183 198 | 1.01 | 1.00-1.02 | 1.93 |
| Uruguay | 238 | 1.61 | 1.34-1.92 | 847 | 0.71 | 0.39-1.13 | 0.90 |
| Venezuela | 119 | 1.52 | 1.13-1.99 | 2 814 | 1.04 | 0.97-1.12 | 0.48 |

$R_0$ , basic reproduction number; $R_t$ , time-dependent reproduction number; 95% CI, 95% confidence interval; 95% CrI, 95% credible interval

## DISCUSSION

We used two measures to estimate the spread of SARS-CoV-2: *R_0_* during the early phase of the outbreak, and *R_t_* to measure the changes of transmissibility over time to identify the effectiveness of public health policies in SA countries. The main results identified that *R_0_* varied across nations, and *R_t_* decreased over time in all countries. The latter, seems to be associated with the timing of implementation of mitigation measures.

Our results differ from previous estimations of *R_0_* and *R_t_*. Li et al. (18) reported an *R_0_* of 2.2 (95% CI: 1.4-3.9) calculated during the first 26 days of the outbreak in Wuhan, China. Moreover, Zhao et al. (19) estimated an *R_0_* between 2.24 (95% CI: 1.96-2.55) and 5.71 (95% CI: 4.24-7.54) withing the first 15 days of the outbreak. We used different parameters to estimate *R_0_*. First, we chose 14 days based on the maximum incubation period of the virus, providing a reasonable window in which transmission can still be considered exponential. This choice renders our model specification consistent with White and Pagano’s method and prevents our *R_0_* values from being underestimated (14). Second, in the absence of contact tracing data, previous studies used the serial interval of other similar respiratory viruses as a proxy for the serial interval of SARS-CoV-2. Li et al. (18) used a mean of 8.4 ±3.8 days from SARS, while Zhao et al. (19) employed a mean of 7.6 ± 3.4 days from MERS; with both studies using a gamma distribution. A study from the CDC identified a lower mean serial interval for SARS-Cov-2—3.96 ± 4.75 days—and proposed a gamma distribution as a plausible model for the serial interval; we adjusted our analysis to this prior.

*R_t_* values from SA countries were lower than those of European nations: *R_t_* estimates by Yuan et al. (20) for Italy, France, Germany, and Spain ranged from 3.10 to 6.56 in an overall 20-day period. In contrast, our study estimated *R_t_* over 90 days after the first case identification; therefore, we stress that the main difference of these outcomes relies on the chosen time interval to estimate *R_t_*. SARS-Cov-2 might have been spreading rapidly during the first 20 days after case detection in Europe, which is reflected in higher *R_t_* values. Another possibility is the longer period that European countries took to implement control measures than SA countries. For example, Italy took over one month to make its first mitigation came into effect (21), which led the virus to spread freely for a longer time in the population.

The reproduction number—*R_0_* or *R_t_*—is a measure that depends on the population mixing (6). Human behavior play a critical role in the transmission of SARS-CoV-2, as person-to-person contact exposes a susceptible person through respiratory droplets from an infected individual (22). Therefore, mitigation measures, such as social distancing or case isolation, are necessary to stop the spread of SARS-CoV-2. Ideal policies should be country-specific, and the overall objective is to reduce *R_0_* (mitigation) or to reach an *R_0_* <1 (suppression). Regardless of the aim, a combination of mitigation and suppression measures is the best strategy to staunch the COVID-19 pandemic (10).

Three countries, Brazil, Chile, and Uruguay opted for a mitigation strategy. Ferguson et al. suggested that the best mitigation strategy is a combination of physical distancing of high-risk groups (elders and patients at risk of severe disease), case isolation, and quarantine (10).

Uruguay implemented these three measures. Brazil and Chile added a stepwise selective population containment, which allows intermittent circulation of SARS-CoV-2, congruent with a mitigation purpose. The remaining SA countries opted for suppression actions. For example, Peru issued a decree to increase the period of the community-wide containment intervention and to fine citizens if they left their home after a specified hour of the day (23). As a result, all nations managed a reduction of *R_t_*.

Here we highlight several explanations for the change between *R_0_* and *R_t_*. First, the implementation of public health measures by some countries within the first week of the outbreak could have lowered *R_0_*. These laws aimed to reduce human contact, which, in turn, reduced the spread of the virus in the community. In contrast, the three countries with the highest *R_0_* in the early phase issued their first mitigation laws ≥14 days from their first cases, which gave SARS-CoV-2 a higher chance of transmissibility among the population. Second, the reproduction number is sensitive to the ability of each country to detect COVID-19 cases (24). A shortage of testing impairs the identification of cases within a community and provides limited data to estimate *R_0_* or *R_t_*, generating unclear information to analyze the effect of mitigation or suppression interventions in a community. In our view, despite international donations of test kits to identify and isolate cases (25, 26), the fragmented healthcare systems and questionable government management of SA countries deter proper development of testing strategy and test distribution for case detection.

Our study has some limitations to address. Although we used national reports of COVID-19 cases, underreporting is likely, and difficult to quantify. True case identification depends on the testing capacity to detect both symptomatic and asymptomatic patients, which is difficult in any country and likely impaired in SA countries. For instance, Venezuela has been under political, socioeconomic, and public health instability before the COVID-19 pandemic, shown through the cessation of publishing of public health statistics since 2016 from their Ministry of Health (27). Moreover, Brazil’s president Jair Bolsonaro downplayed COVID-19 regarding it as a benign disease and through avoiding prompt action to address the pandemic (28). These instances may have impaired proper case detection and led to a suppression of information as previous reports suggest (28, 29).

Second, we did not calculate the case-fatality ratio. During the current course of the pandemic in SA countries, the estimation of this metric is biased for underreporting of cases and time lag between the notification of cases and deaths; this analysis is better suited for a post-pandemic period (30, 31). Third, there are issues arising from the model specification for serial interval. For instance, we assumed the same serial interval distribution for all countries at all points in time, even though these should be space- and time-dependent, as serial interval distributions vary throughout an epidemic or pandemic (24). Also, the CDC serial interval model only considers positive serial times, and hence censors all serial interval observations in which a secondary case manifests symptoms before the primary case does. Per their findings, about 12.6% of secondary cases exhibit clinical symptoms before a primary case (e.g. an infected person presenting symptoms before the infector); given how sensitive *R_0_* estimates are to the serial interval distribution, our results should be updated as better model specifications of this variable are elucidated.

Our findings suggest a positive, yet insufficient, impact of the mitigation and suppression measures in SA nations to reduce the spread of SARS-CoV-2. Despite the fragmented health system of most of these countries, the different combination of control measures adopted managed to reduce *R_t_*, albeit to different levels. The difference of *R_0_* in the early phase of the outbreak is probably due to a combination of a shortage of testing and the idiosyncrasies of each country’s public health system. However, *R_t_* values above one during the study period suggest that South America is still far from containing the spread of COVID-19.

## Data Availability

Data is freely available

https://www.ecdc.europa.eu/en/publications-data/download-todays-data-geographic-distribution-covid-19-cases-worldwide

## ACKNOWLEDGMENTS

We thank Dr. Sanz-Anquela JM., for the advice given in this work and Lopez-Abente G., for encouraging us to conduct this study, and Chloé Mauvais for her insightful comments and for proofreading the manuscript.

